# Association of “Metabolic Dysfunction-Associated Steatotic Pancreas Disease” (MASPD) with Insulin Resistance

**DOI:** 10.1101/2024.01.13.24301274

**Authors:** Luís Jesuino de Oliveira Andrade, Gabriela Correia Matos de Oliveira, Alcina Maria Vinhaes Bittencourt, Guilherme Peixoto Nascimento, Gustavo Magno Baptista, Catharina Peixoto Silva, Luís Matos de Oliveira

## Abstract

**Introduction:** “Metabolic Dysfunction-Associated Steatotic Pancreas Disease” (MASPD) is not yet a term or condition described in the medical literature. The MASPD is a relatively new and emerging condition that has garnered significant attention in the field of metabolic disorders.

**Objective:** to investigate the association between MASPD and IR and explore the potential mechanisms that may contribute to this relationship.

**Materials and Methods:** This cross-sectional study involved 157 participants diagnosed with MASPD based on ultrasonography criteria. Baseline demographic data were collected, including age, gender, and body mass index. Serum levels of fasting glucose, insulin, lipid profile (including total cholesterol, triglycerides, high-density lipoprotein cholesterol, and low-density lipoprotein cholesterol), glycated hemoglobin and insulin were measured using standardized laboratory techniques. Abdominal ultrasonography was performed on all participants using convex transducer (frequency range, 3,5 MHz) by experienced radiologist blinded to the clinical data. The association between MASPD and IR was assessed using logistic regression analysis, adjusting for potential confounders. Statistical significance was set at a p-value of less than 0.05.

**Results:** The logistic regression analysis was performed to verify whether IR was a risk factor for MASPD. After adjusting for gender and age, the results demonstrate a significant correlation between MDASPD and markers of IR. TyG index: OR (95% IC) 5.72 (1.90 – 16.00), *P* 0.021, and HOMA –IR: OR (95% IC) 6.20 (2.1 – 22.00) *P* 0.037.

**Conclusion:** This study presents the first description of MDASPD and its association with IR indices. Our findings demonstrate a significant correlation between MDASPD and markers of IR. These results suggest that MDASPD may contribute to the development of insulin resistance and further highlight the importance of pancreatic health in metabolic disorders.

## INTRODUCTION

“Metabolic Dysfunction-Associated Steatotic Pancreas Disease” (MASPD) is not yet a term or condition described in the medical literature. The MASPD is a relatively new and emerging condition that has garnered significant attention in the field of metabolic disorders. This disease is characterized by pancreatic fat infiltration in the absence of chronic pancreatitis.^1^ Although the exact pathogenesis of MASPD remains unclear, there is growing evidence suggesting an association between MASPD and insulin resistance (IR).^2,3^

IR is a key feature of various metabolic disorders, including type 2 diabetes mellitus, metabolic syndrome, and non-alcoholic fatty liver disease. It is characterized by impaired insulin signaling and reduced glucose uptake in insulin target tissues such.^4^

The prevalence of MASPD appears to be on the rise, mirroring the global epidemic of obesity and type 2 diabetes mellitus.^5^ As a consequence, understanding the association between MASPD and IR has become increasingly important for a better comprehension of the pathophysiology of both conditions. Moreover, identifying this relationship may have clinical implications in the prevention and management of MASPD and its associated metabolic comorbidities

The pancreas plays a important role in glucose homeostasis through the secretion of insulin and other hormones involved in the regulation of glucose metabolism. Any alteration in pancreatic function can have profound effects on glucose control and metabolic processes. Emerging evidence suggests that pancreatic steatosis, a hallmark feature of MASPD, may disrupt the normal pancreatic function, leading to impaired insulin secretion.^6,7^

There is a growing body of evidence suggesting a bidirectional relationship between MASPD and IR.^8^ However, the precise mechanisms underlying this association remain incompletely understood, necessitating further investigation.

Despite the growing interest in understanding the relationship between MASPD and IR, there is a paucity of research investigating the precise mechanisms underlying this association. Moreover, the clinical implications of this association remain poorly defined. Thus, the objective of this research is to investigate the association between MASPD and IR and explore the potential mechanisms that may contribute to this relationship.

## MATERIALS AND METHODS

### Study Design and Participants

This cross-sectional study involved 157 participants diagnosed with MASPD based on ultrasonography criteria. The study protocol was approved by the Ethics Committee.

### Clinical and Laboratory Assessments

Baseline demographic data were collected, including age, gender, body mass index. Fasting blood samples were obtained from each participant for laboratory assessments. Serum levels of fasting glucose, insulin, lipid profile (including total cholesterol, triglycerides, high-density lipoprotein cholesterol, and low-density lipoprotein cholesterol), glycated hemoglobin and insulin were measured using standardized laboratory techniques.

### Assessment of Insulin Resistance

IR was assessed using the TyG index and HOMA-IR. The TyG index was determined as Ln[fasting triglycerides (mg/dL) fasting glucose (mg/dL)/2], and it is expressed on a logarithmic scale.^9^ The benchmark for identifying IR is established at a TyG index value of 4.49. The HOMA-IR index was derived using the following formula: (Fasting insulin in betaU/mL × fasting glucose in mg/dL) / 415.^10^ In accordance with earlier research, an HOMA-IR index ≥ 3.4 indicates IR, which represents the optimal threshold for predicting the development of diabetes mellitus and aligns with the hyperglycemic-hyperinsulinemic clamp method.

### Pancreatic Imaging

Abdominal ultrasonography was performed on all participants using convex transducer (frequency range, 3,5 MHz) by experienced radiologists blinded to the clinical data. Pancreatic steatosis was assessed based on the presence and severity of hyperechogenicity in comparison to the liver parenchyma. The severity of pancreatic steatosis was graded as absent, mild (Grade I), moderate (Grade II), or severe (Grade III), according to predefined criteria.

### Statistical Analysis

Descriptive statistics were presented as mean ± standard deviation (SD) or median (interquartile range) for continuous variables, and percentages for categorical variables. The association between MASPD and IR was assessed using logistic regression analysis, adjusting for potential confounders. Statistical significance was set at a *P*-value of less than 0.05.

### Ethical Considerations

This study was conducted in accordance with the principles outlined in the Declaration of Helsinki. The study protocol The National Commission for Research Ethics (CONEP - Brazil) approved the project (registry number: 2.464.513) was approved by The National Commission for Research Ethics (CONEP - Brazil), and written informed consent was obtained from all participants prior to their inclusion in the study. Confidentiality of personal information was strictly maintained throughout the study, and all data were analyzed and reported in an aggregated and anonymized manner.

## RESULTS

A comprehensive assessment was conducted on a total of 157 specimens, comprising of 126 (63.0%) females and 74 (37.0%) males, who had an average age of 46.56 ± 18.98 years. IR was detected in 64 individuals (32.0%) based on the HOMA-IR index and in 132 individuals (66.0%) based on the TyG index. The HOMA-IR and TyG index demonstrated distinct patterns within the samples under evaluation. The summarized demographic features can be found in Table 1 and Table 2.

**Table 1.** Demographic and biochemical characteristics.

| **Gender** | **Male: 58 (36.9%) Female: 99 (63.1%)** |
| --- | --- |
| **Age (years)** | 52.29 ± 17.91 |
| **HBA1c** | 5,78 ± 0.91 |
| **Insulin** | 12.06 ± 9.12 |
| **Fasting glucose** | 106.90 ± 23.24 |
| **Average blood glucose estimated by HBA1C** | 119.26 ± 25.99 |
| **Triglycerides** | 124.84 ± 66.63 |
| **HOMA-IR** | 3.13 ± 2.74 |
| **TyG index** | 4.64 ± 0.30 |

**Table 2.**
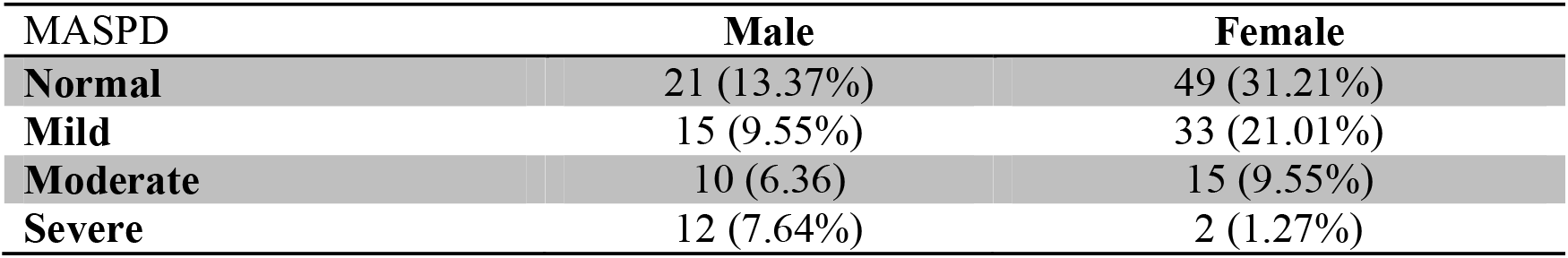
Parameters of graded MASPD.

In our study, we employed the ultrasonographic method to evaluate the degree of pancreatic fat infiltration and utilized the UFPS to classify the severity of pancreatic fat infiltration.

The logistic regression analysis was performed to verify whether IR was a risk factor for MASPD. After adjusting for gender and age, the results are shown in Table 3.

**Table 3.** Logistic analysis of MASPD risk factors.

| Variables | <sup>a</sup>OR (95% CI) | *P* value |
| --- | --- | --- |
| **TyG index** | 5.72 (1.90 – 16.00) | 0.021 |
| **HOMA –IR** | 6.20 (2.1 – 22.00) | 0.037 |

<sup>a</sup>Adjusted for gender and age

## DISCUSSION

In this study, we demonstrated that the TyG index and HOMA-IR levels serve as significant risk factors for the development of MASPD. Our analysis revealed a strong correlation between these metabolic indices and the pathogenesis of the disease, providing critical insights into the underlying mechanisms and potential therapeutic targets for this increasingly prevalent condition.

The MASPD is a complex disorder characterized by pancreatic steatosis.^11^ Despite its clinical importance, the risk factors contributing to MASPD remain poorly understood. In recent years, the TyG index and HOMA-IR have emerged as potential predictors of metabolic dysfunction and IR in various metabolic diseases.^12^ Building upon this knowledge, this study aimed to investigate the association between the TyG index, HOMA-IR levels, and the risk of developing MASPD. By elucidating the role of these metabolic markers in MASPD pathogenesis, our findings may provide a foundation for early detection, risk stratification, and targeted interventions to mitigate the progression and complications of this pancreatic disorder.

The assessment of IR is crucial in clinical practice, and while the HOMA-IR index has been widely used as a reliable marker, and the TyG index has emerged as a promising alternative.^13,14^ HOMA-IR, calculated from fasting insulin and glucose levels, provides an established measure of insulin sensitivity, but it may underestimate IR in certain populations due to its reliance on fasting insulin levels.^15^ In contrast, the TyG index, derived from fasting triglycerides and glucose concentrations, offers a simple and practical surrogate for IR assessment, demonstrating associations with metabolic risk.^16^ However, both indices have limitations, such as variability in cutoff values and sensitivity to changes in lipid and glycemic profiles, warranting cautious interpretation in clinical settings.^17^ Overall, the combined use of HOMA-IR and TyG index enhances the precision of IR evaluation.^18^

The relationship between the TyG index and HOMA-IR with MASPD has been underexplored, despite evidence of the role of these indices in the pathophysiology of metabolic disorders such as NASH. Studies have shown that both an increase in the TyG index and HOMA-IR are associated with the development of NASH, suggesting the importance of these markers in the assessment of hepatic steatosis.^19,20^ However, few studies have investigated the potential association between these indices and MASPD. Therefore, our research aimed to evaluate the relationship between the TyG and HOMA-IR indices with the degrees of MASPD, demonstrating a significant association between them.

Ultrasonographic diagnosis of pancreatic fat infiltration involves the assessment of echogenicity and texture of the pancreatic parenchyma. Various grading systems have been proposed to classify the severity of pancreatic steatosis based on ultrasound findings. The ultrasonographic fatty pancreas score (UFPS) assesses the degree of pancreatic fat infiltration based on the echogenicity of the pancreas compared to the liver and kidney[19].^21^ In our study, we employed the ultrasonographic method to evaluate the degree of pancreatic fat infiltration and utilized the UFPS to classify the severity of pancreatic fat infiltration.

Thus, we are providing the first description of the potential mechanisms linking insulin resistance and MDASPD, along with the concurrent elevation of triglycerides, glucose, and insulin levels. IR, characterized by impaired insulin signaling and decreased glucose uptake in target tissues, may lead to increased lipolysis in adipose tissue, resulting in elevated circulating free fatty acids.^22^ These free fatty acids are then taken up by the liver and pancreas, leading to increased triglyceride accumulation within these organs.^23,24^ Simultaneously, IR disrupts the normal regulation of glucose uptake and metabolism, resulting in hyperglycemia and compensatory hyperinsulinemia. The combination of elevated triglycerides, glucose, and insulin levels contributes to an environment conducive to the development of MDASPD.

## CONCLUSION

This study presents the first description of MDASPD and its association with insulin resistance indices. Our findings demonstrate a significant correlation between MDASPD and markers of IR. These results suggest that MDASPD may contribute to the development of IR and further highlight the importance of pancreatic health in metabolic disorders. Further investigations are warranted to elucidate the underlying mechanisms and potential therapeutic strategies targeting MDASPD and its impact on insulin resistance.

## Data Availability

All data produced in the present study are available upon reasonable request to the authors

## References

1. Mahyoub MA, Elhoumed M, Maqul AH, Almezgagi M, Abbas M, Jiao Y, et al. Fatty infiltration of the pancreas: a systematic concept analysis. Front Med (Lausanne). 2023;10:1227188.

2. Wen Y, Chen C, Kong X, Xia Z, Kong W, Si K, et al. Pancreatic fat infiltration, β-cell function and insulin resistance: A study of the young patients with obesity. Diabetes Res Clin Pract. 2022;187:109860.

3. Larsen MO, Juhl CB, Pørksen N, Gotfredsen CF, Carr RD, Ribel U, et al. Beta-cell function and islet morphology in normal, obese, and obese beta-cell mass-reduced Göttingen minipigs. Am J Physiol Endocrinol Metab. 2005;288(2):E412–21.

4. Bonora E, Kiechl S, Willeit J, Oberhollenzer F, Egger G, Targher G, et al. Prevalence of insulin resistance in metabolic disorders: the Bruneck Study. Diabetes. 1998;47(10):1643–9.

5. Guglielmi V, Sbraccia P. Type 2 diabetes: Does pancreatic fat really matter? Diabetes Metab Res Rev. 2018;34(2).

6. Ramkissoon R, Gardner TB. Pancreatic Steatosis: An Emerging Clinical Entity. Am J Gastroenterol. 2019;114(11):1726–1734.

7. Begovatz P, Koliaki C, Weber K, Strassburger K, Nowotny B, Nowotny P, et al, Pancreatic adipose tissue infiltration, parenchymal steatosis and beta cell function in humans. Diabetologia. 2015;58(7):1646–55.

8. Wagner R, Eckstein SS, Yamazaki H, Gerst F, Machann J, Jaghutriz BA, et al. Metabolic implications of pancreatic fat accumulation. Nat Rev Endocrinol. 2022;18(1):43–54.

9. Simental-Mendía LE, Rodríguez-Morán M, Guerrero-Romero F. The product of fasting glucose and triglycerides as surrogate for identifying insulin resistance in apparently healthy subjects. Metab Syndr Relat Disord. 2008;6(4):299–304.

10. Matthews DR, Hosker JP, Rudenski AS, Naylor BA, Treacher DF, Turner RC. Homeostasis model assessment: insulin resistance and beta-cell function from fasting plasma glucose and insulin concentrations in man. Diabetologia. 1985;28(7):412–9.

11. Paul J, Shihaz AVH.PANCREATIC STEATOSIS: A NEW DIAGNOSIS AND THERAPEUTIC CHALLENGE IN GASTROENTEROLOGY. Arq Gastroenterol. 2020;57(2):216–220.

12. Zhao X, An X, Yang C, Sun W, Ji H, Lian F. The crucial role and mechanism of insulin resistance in metabolic disease.Front Endocrinol (Lausanne). 2023;14:11492 39.

13. Gayoso-Diz P, Otero-Gonzalez A, Rodriguez-Alvarez MX, Gude F, Cadarso-Suarez C, García F, et al. Insulin resistance index (HOMA-IR) levels in a general adult population: curves percentile by gender and age. The EPIRCE study.Diabetes Res Clin Pract. 2011;94(1):146–55.

14. Tahapary DL, Pratisthita LB, Fitri NA, Marcella C, Wafa S, Kurniawan F, et al. Challenges in the diagnosis of insulin resistance: Focusing on the role of HOMA-IR and Tryglyceride/glucose index. Diabetes Metab Syndr. 2022;16(8):102581.

15. So A, Sakaguchi K, Okada Y, Morita Y, Yamada T, Miura H, et al. Relation between HOMA-IR and insulin sensitivity index determined by hyperinsulinemic-euglycemic clamp analysis during treatment with a sodium-glucose cotransporter 2 inhibitor. Endocr J. 2020;67(5):501–507.

16. Ramdas Nayak VK, Satheesh P, Shenoy MT, Kalra S. Triglyceride Glucose (TyG) Index: A surrogate biomarker of insulin resistance. J Pak Med Assoc. 2022;72(5):986–988.

17. Fernandes AC, Marinho AR, Lopes C, Ramos E. Dietary glycemic load and its association with glucose metabolism and lipid profile in young adults. Nutr Metab Cardiovasc Dis. 2022;32(1):125–133

18. Son DH, Lee HS, Lee YJ, Lee JH, Han JH. Comparison of triglyceride-glucose index and HOMA-IR for predicting prevalence and incidence of metabolic syndrome. Nutr Metab Cardiovasc Dis. 2022;32(3):596–604.

19. Xue Y, Xu J, Li M, Gao Y. Potential screening indicators for early diagnosis of NAFLD/MAFLD and liver fibrosis: Triglyceride glucose index-related parameters. Front Endocrinol (Lausanne). 2022;13:951689.

20. Moriyama K, Inoue N, Imai J, Masuda Y, Yamada C, Kishimoto N, et al. Prediction and Validation of Metabolic Dysfunction-Associated Fatty Liver Disease Using Insulin Resistance-Related Indices in the Japanese Population. Metab Syndr Relat Disord. 2023;21(9):489–496.

21. Romana BS, Chela H, Dailey FE, Nassir F, Tahan V. Non-Alcoholic Fatty Pancreas Disease (NAFPD): A Silent Spectator or the Fifth Component of Metabolic Syndrome? A Literature Review. Endocr Metab Immune Disord Drug Targets. 2018;18(6):547–554.

22. Kraegen EW, Cooney GJ, Ye J, Thompson AL. Triglycerides, fatty acids and insulin resistance--hyperinsulinemia. Exp Clin Endocrinol Diabetes. 2001;109(4):S516–26.

23. Vergani L. Fatty Acids and Effects on In Vitro and In Vivo Models of Liver Steatosis. Curr Med Chem. 2019;26(19):3439–3456.

24. Phillips AE, Wilson AS, Greer PJ, Hinton A, Culp S, Paragomi P, et al. Relationship of circulating levels of long-chain fatty acids to persistent organ failure in acute pancreatitis. Am J Physiol Gastrointest Liver Physiol. 2023;325(3):G279–G285.

